# Client-perpetrated gender-based violence among female sex workers in post-conflict Gulu district, Northern Uganda: a cross-sectional study

**DOI:** 10.1101/2020.11.02.20224527

**Authors:** Simple Ouma, Rawlance Ndejjo, Catherine Abbo, Nazarius Mbona Tumwesigye

## Abstract

**Background:** Gender-based violence (GBV) among female sex workers (FSWs) negatively affects their mental wellbeing and sexual and reproductive health and rights. This study aimed to determine the prevalence and associated factors of client-perpetrated GBV among FSWs in post-conflict Gulu district, Northern Uganda.

**Methods:** A cross-sectional study was conducted among 300 FSWs aged 18+ years who were operating in Gulu district. Participants were selected using simple random sampling and interviewed between March and June 2020. Pre-tested semi-structured questionnaires were utilized to collect information on socio-demographic and sex-work-related characteristics, alcohol and illicit drug use, HIV status, and exposure to client-perpetrated GBV. Data were entered into EPI INFO 7 and analyzed using logistic regression with the aid of STATA 14.0.

**Results:** Sixty one percent (61.0%) of the participants reported client-perpetrated GBV. The most common forms of GBV in this population were economic (58.7%) and emotional (52.0%) violence. Meanwhile, sexual violence (21.0%) was the least common form of GBV among the study participants. At multivariate level; street-based sex work (aOR=9.66, 95%CI: 2.78-33.5), mobile sex work (aOR=3.21, 95%CI: 1.83-5.64), HIV-positive status (aOR=1.90, 95%CI: 1.09-3.31), and low monthly income below 50^th^ percentile {<200,000 UGX}(aOR= 2.26, 95% CI: 1.18-4.30) remained independently associated with client-perpetrated GBV.

**Conclusion:** Our findings revealed a high prevalence of client-perpetrated GBV driven by street-based sex work, sex work-related mobility, HIV-positive status, and low income. Therefore, interventions to address client-perpetrated GBV should target FSWs who are street-based, HIV-positive, mobile, and with low income.

**Strengths and limitations of this study:**

▪ Unlike most previous studies among the FSWs that used the non-probability methods of participants’ selection, we utilized a simple random sampling technique to select our sample. Thus, our study participants were representative of the FSWs.
▪ However, the information collected may have been influenced by recall bias since we asked FSWs about their personal experience of client-perpetrated GBV.
▪ In addition, we implored some sensitive information relating to sex work that might have been difficult to answer which might have resulted into information bias.

I, the Submitting Author has the right to grant and does grant on behalf of all authors of the Work (as defined in the below author licence), an exclusive licence and/or a non-exclusive licence for contributions from authors who are: i) UK Crown employees; ii) where BMJ has agreed a CC-BY licence shall apply, and/or iii) in accordance with the terms applicable for US Federal Government officers or employees acting as part of their official duties; on a worldwide, perpetual, irrevocable, royalty-free basis to BMJ Publishing Group Ltd (“BMJ”) its licensees and where the relevant Journal is co-owned by BMJ to the co-owners of the Journal, to publish the Work in this journal and any other BMJ products and to exploit all rights, as set out in our [licence](https://authors.bmj.com/wp-content/uploads/2018/11/BMJ_Journals_Combined_Author_Licence_2018.pdf).

The Submitting Author accepts and understands that any supply made under these terms is made by BMJ to the Submitting Author unless you are acting as an employee on behalf of your employer or a postgraduate student of an affiliated institution which is paying any applicable article publishing charge (“APC”) for Open Access articles. Where the Submitting Author wishes to make the Work available on an Open Access basis (and intends to pay the relevant APC), the terms of reuse of such Open Access shall be governed by a Creative Commons licence – details of these licences and which [Creative Commons](http://creativecommons.org/) licence will apply to this Work are set out in our licence referred to above.

Other than as permitted in any relevant BMJ Author’s Self Archiving Policies, I confirm this Work has not been accepted for publication elsewhere, is not being considered for publication elsewhere and does not duplicate material already published. I confirm all authors consent to publication of this Work and authorise the granting of this licence.

## Introduction

The United Nations defined violence against women as “any act of GBV that results in or is likely to result in, physical, sexual or psychological harm or suffering to women, including threats of such acts, coercion or arbitrary deprivation of liberty, whether occurring in public or private life”(1). Survivors of GBV often fail to achieve legal, social, political, and economic equality in society (1). A previous study indicated that the majority of FSWs were affected by both intimate and nonintimate partner violence (2). Moreover, the management of GBV in this vulnerable population remains suboptimal since FSWs have poor access to health care services for fear of victimization by the health care providers (3,4). Besides, the impacts of political conflict might have worsened the GBV among FSWs operating in settings like Gulu district yet client-perpetrated GBV in post-conflict settings remain understudied. Additionally, sex work is illegal and not recognized as a form of employment in Uganda (5). This illegality of sex work may have led to an increase in workplace violence among FSWs in the country since the law does not protect them.

Currently, there is a growing body of literature on risk factors of GBV among FSWs. One such evidence suggests that HIV infection increases the risk of GBV (6). However, the relationship between GBV and HIV is bidirectional since survivors of GBV also tend to have a greater risk of acquiring HIV infection (7,8). Secondly, compared to FSWs who provide sexual services from one locality, mobile sex workers are more likely to get exposed to client-perpetrated GBV (9,10). Additionally, a systematic review revealed that both alcohol and illicit drug use increase intimate and nonintimate partner GBV among FSWs (11). Moreover, if left untreated, GBV could lead to poverty, alcohol abuse, reduction in condom use consistency (12,13), decrease in condom self-efficacy with clients (14), reduction in the uptake of sexual and reproductive health services (15), and mental disorders like depression (16–19). Thus, addressing client-perpetrated GBV among the FSWs is of public health importance and is very critical for the improvement of mental wellbeing and promotion of sexual reproductive health and rights in this vulnerable and marginalized population.

In order to prevent and manage GBV among FSWs in the country, understanding the epidemiology of GBV in this population is very crucial since findings provide the evidence needed for the development of evidence-based and context-specific interventions. Yet the prevalence and risk factors of client-perpetrated GBV among FSWs living and working in post-conflict settings like Gulu district have not been well explored. The specific objectives of this study were 1) to determine the prevalence of client-perpetrated GBV among FSWs in post-conflict Gulu district, Uganda and 2) to determine factors associated with client-perpetrated GBV among FSWs in the post-conflict Gulu district, Uganda.

## Methods

### Study setting, design, and population

We conducted a study among FSWs in the post-conflict Gulu district, Northern Uganda. FSWs are considered a key population and are targeted for HIV prevention because of the high burden of HIV among them (20). People in Gulu districts are still recovering from the more than 20 years of Lord Resistant Army (LRA) rebellion that devastated their social and economic livelihoods. More than 80% of the inhabitants of the district practice subsistence farming (21) and an estimated 1425 FSWs operate in Gulu district (22), the majority of whom operate in Gulu municipality. Unpublished program data show that more than 1300 fully mapped-out FSWs receive HIV treatment and/or preventive services from TASO Gulu. We conducted a cross-sectional study among FSWs aged 18+ years who were operating in the district.

### Sample size and sampling

We collected data among 300 FSWs between March and June 2020. The sample size was determined using the Cochran [1963,1975](23) formula: n_0_ = Z^2^pq/e^2^. This work was part of a project that examined the epidemiology of depression among FSWs in Gulu district on which the sample size determination was based and the calculated sample size was 303. The sample size was adjusted 380 for work-related mobility (10%) and non-response (10%). We utilized a simple random sampling technique because we had an up-to-date database of FSWs at TASO Gulu. However, with the help of the peers of FSWs, we had to rapidly update the existing database to include any missing FSW in the database.

### Data collection

Data were collected through face-to-face interviews using a pre-tested semi-structured questionnaire developed in English and translated into Acholi language (*Luo*), the most widely spoken local language in the district. The first author and a trained female research assistant collected data in either Acholi or English language depending on the participant’s literacy level and preference. Independent variables included socio-demographic characteristics like age, education, religion, and marital status, sex work-related characteristics like duration of sex work, average monthly income, place of sex work, and sex work-related mobility, alcohol use, illicit drug use, and HIV status. To determine intra-regional sex work-related mobility, we asked the participants whether they provided sexual services in an urban or a rural setting only, or both urban and rural settings. We considered FSWs who provided sexual services in both rural and urban settings to be mobile. Our dependent variable was client-perpetrated GBV. We asked each participant whether her client(s) ever: refused to pay for sexual services (economic), verbally abused (insulted) her (emotional), physically abused/beat her (physical), or forced her to have sex/raped her (sexual violence). Reporting either economic, emotional, physical, or sexual violence by male clients was considered client-perpetrated GBV.

### Data management and statistical analysis

Data were entered and cleaned in EPI INFO 7 and then exported to STATA 14.0 for analyses. We described univariate analyses using frequencies with their corresponding proportions for categorical variables and means with corresponding standard deviations for continuous variables. We conducted both bivariate and multivariate logistic regression analyses to examine the associations of the independent factors with client-perpetrated GBV. We presented the results of bivariate analyses using unadjusted odds ratios (UOR) with corresponding confidence intervals (CI) and p-value. Then, using all the significant (p<0.20) independent variables at bivariate analysis, we constructed a multivariate logistic regression model to identify factors associated with client-initiated GBV. Thus, controlling for age, marital status, average monthly income, and illicit drug use, we entered all eligible independent variables into the multivariate logistic regression model at the beginning stage of model building. We utilized the backward elimination method and sequentially removed each factor with the least significant p-value while testing the model fit using the goodness-of-fit test until we obtained the best fit model. Results from multivariable analyses were presented using adjusted odds ratios (AOR) with corresponding 95% CIs and p-value.

After fitting the adjusted regression model, we conducted sensitivity analysis and regression diagnostics tests on the best fitted model. Specifically, we investigated the predictive power of the model using the sensitivity and specificity analyses, and performed the linktest to check for specification error.

### Patient and Public Involvement

We involved the patients and the public in the dissemination plans of our research; findings were shared with the relevant authorities in the district local government and the non-governmental organizations providing health care services to the key population in the region. It is worth noting that the complex nature of FSWs made it very difficult to actively involve the patients or the public in the design, or conduct, or reporting plans of our research. However, the first author’s personal experience and interactions with the FSWs and their peers, during his more than three years of providing HIV and sexual-reproductive health services to the key population that include the FSWs, informed the design and conducts of the study. Additionally, the peers of FSWs were involved in the identification and location of the selected participants.

## Results

### Socio-demographic and sex work-related characteristics of study participants

The mean age of the study participants was 26.4 years (SD ± 6, Range = 18-50 years) majority (40%) of whom were below 25 years old. Additionally, 62.0% of the participants had no or primary education, 51.3% had migrated to Gulu district, only 14.3% were married/cohabiting, and 60.7% were Catholics. The majority (39.7%) of the participants had been sex workers for 3-5 years, 63.9% had sex work as their main source of income and, 43.5% earned below the 50^th^ percentile of the average monthly income of the FSWs (200,000 UGX (60$)) (Table 1).

**Table 1:** Socio-demographic and sex work-related characteristics of study participants.

|  |  |  |
| --- | --- | --- |
| Characteristic | Number (N) | Percent (%) |
| Age (completed years) |  |  |
| <25 | 120 | 40.0 |
| 25-29 | 85 | 28.3 |
| 30+ | 95 | 31.7 |
| Education |  |  |
| ≤ Primary level | 186 | 62.0 |
| ≥ Secondary level | 114 | 38.0 |
| District of Origin |  |  |
| Gulu | 146 | 48.7 |
| Others | 154 | 51.3 |
| Married/Cohabiting |  |  |
| No | 257 | 85.7 |
| Yes | 43 | 14.3 |
| Religion |  |  |
| None/others | 36 | 12.0 |
| Catholic | 182 | 60.7 |
| Protestant | 48 | 16.0 |
| Born Again | 34 | 11.3 |
| Years of sex work |  |  |
| ≤ 2 | 108 | 36.0 |
| 3-5 | 119 | 39.7 |
| >5 | 73 | 24.3 |
| Sex work is the main source of income |  |  |
| No | 108 | 36.1 |
| Yes | 191 | 63.9 |
| Monthly income (UGX) |  |  |
| ≥300,000 | 114 | 38.1 |
| 200,000-<300,000 | 55 | 18.4 |
| <200,000 | 130 | 43.5 |
| Practice mobile sex work |  |  |
| No | 161 | 53.7 |
| Yes | 139 | 46.3 |
| Use illicit drug |  |  |
| No | 216 | 72.0 |
| Yes | 84 | 28.0 |
| Consume alcohol within the Week of the interview |  |  |
| No | 107 | 36.3 |
| Yes | 188 | 63.7 |
| Frequency of alcohol consumption |  |  |
| Never | 79 | 26.5 |
| Occasional | 52 | 17.5 |
| Daily | 167 | 56.0 |
| Reported to be HIV positive |  |  |
| No | 173 | 57.9 |
| Yes | 126 | 42.1 |

### Prevalence of client-perpetrated GBV among FSWs in Gulu district

During sex work, 61.0% of the participants experienced at least one incidence of client-perpetrated GBV. The most common forms of GBV reported were economic (58.7%) and emotional (52.0%) violence and sexual violence (21.0%) was the least common form of GBV. One-fifth (19.0%) of FSWs reported experiencing all the forms of GBV (Table 2).

**Table 2:** Prevalence and forms of client-perpetrated GBV among study participants.

| Characteristics | Numbers (n =300) | Percent (%) |
| --- | --- | --- |
| Experienced client-perpetrated GBV |  |  |
| No | 117 | 39.0 |
| Yes | 183 | 61.0 |
| Client-perpetrated GBV experienced |  |  |
| Economic violence | 176 | 58.7 |
| Emotional violence | 156 | 52.0 |
| Physically violence | 145 | 48.3 |
| Sexual violence | 63 | 21.0 |
| All forms of violence | 57 | 19.0 |

### Predictors of client-perpetrated GBV among female sex workers

At bivariate level, participants who reported client-perpetrated GBV were more likely to be based on the street (uOR=12.0, 95% CI: 3.64-39.8, *p<0*.*001*), club (uOR= 3.16, 95% CI: 1.66-6.00, *p<0*.*001*), brothel (uOR=2.91, 95% CI: 1.80-4.70, *p<0*.*001*), mobile (uOR=3.99, 95% CI: 2.41-6.62, *p<0*.*001*), or the bar (uOR: 2.16, 95% CI: 1.29-3.60, *p=0*.*003*). The other associated factors with client-perpetrated GBV were illicit drug use (uOR= 2.21, 95% CI: 1.27-3.86, *p=0*.*005*), age 25-29 years (uOR=2.05, 95% CI: 1.14-3.68, *p*=0.016), and being HIV positive (uOR: 1.72, 95% CI: 1.07-2.79, *p=0*.*026*).

Results of multivariate logistic regression analysis showed that factors that remained independently associated with client-perpetrated GBV were street-based sex work (aOR=9.66, 95%CI: 2.78-33.5), sex work-related mobility (aOR=3.21, 95%CI: 1.83-5.64), being HIV-positive (aOR=1.90, 95%CI: 1.09-3.31), and low monthly income compared with highest monthly income category (aOR= 2.26, 95% CI: 1.18-4.30). Regression diagnostic tests showed that the adjusted model had; good predictive power (0.77), non-significant goodness-of-fit test outcome (p=0.97), and no specification error (linktest hatsq, p=0.38) (Table *3*).

**Table 3:**
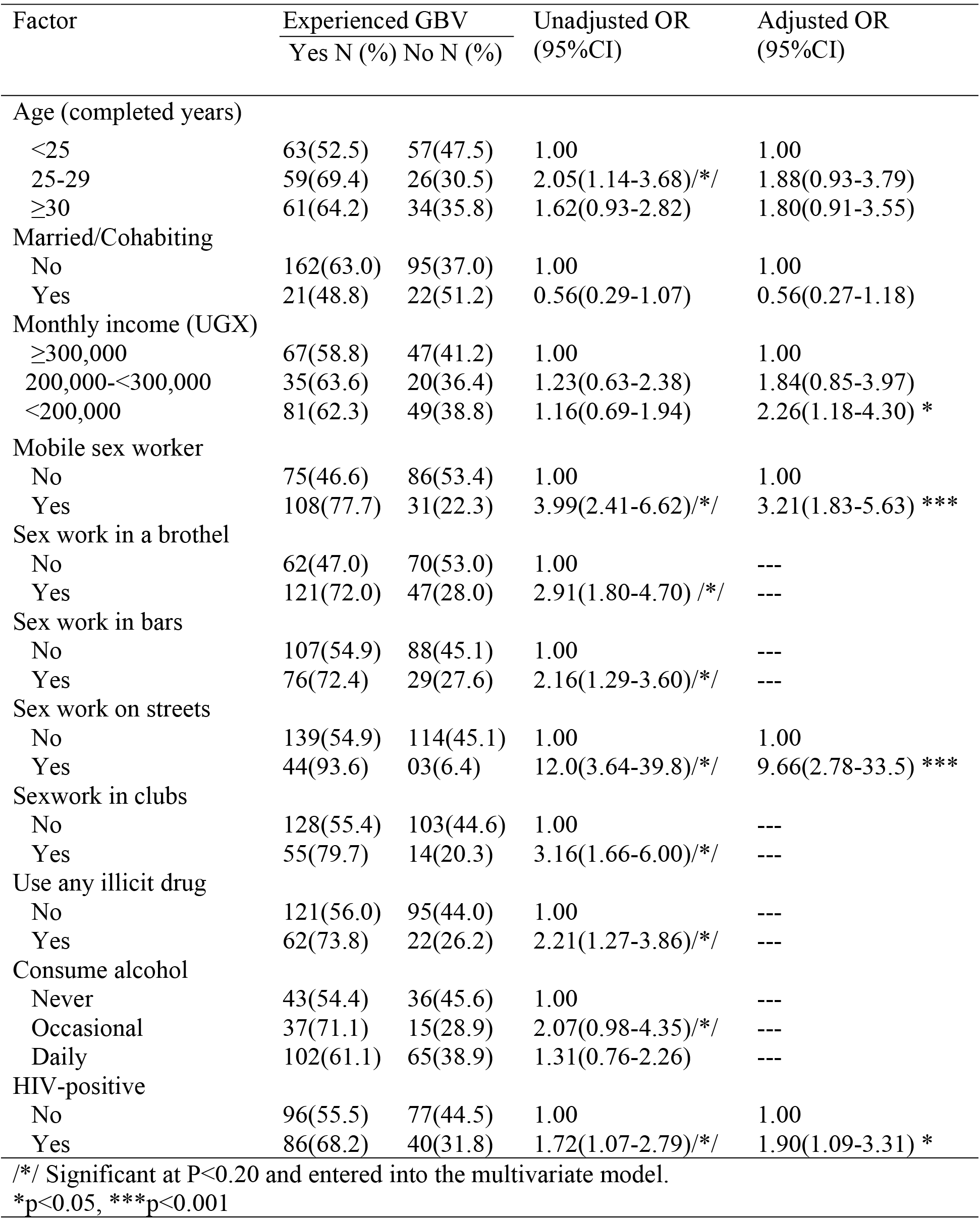
Factors associated with client-perpetrated GBV among FSWs.

## Discussion

Our findings revealed a high prevalence of client-perpetrated GBV among FSWs with the majority reporting emotional and economic violence. Further analysis indicated that the odds of client-perpetrated gender-based violence increased among FSWs who were street-based, mobile, illicit drug users, HIV-positive, and had lower monthly income.

There is a high prevalence of client-perpetrated GBV among FSWs in Gulu district (61%) with the most common forms of violence reported being emotional and economic. This is an increase beyond the previous magnitude of client-perpetrated GBV (50%) among FSWs in Gulu district (24). Cautions should be taken while interpreting these two results since the two studies had some slight measurement variation. In the current study, we measured client-perpetrated GBV for the entire period of sex work as opposed to the previous report that examined client-perpetrated GBV only during the last six months. However, the prevailing magnitude of client-perpetrated GBV is lower than the 82% prevalence of GBV reported among FSWs in Kampala city, Uganda’s Capital (25). We posit that FSWs in Gulu district recorded less client-perpetrated GBV than their Kampala counterparts. Almost all participants in the Kampala study (95%) depended on sex work as their main source of income compared to 63.9% of the current participants who depended on sex work as their main source of income. Results from our study revealed a high magnitude of client-perpetrated GBV among FSWs in the post-conflict Gulu district and calls for urgent public health interventions, especially in the light of the close relationship between GBV and HIV infection (26).

Our findings indicated that street-based FSWs were more likely to experience client-perpetrated GBV. The available literature is in agreement with the current finding showing that FSWs who work in riskier settings like street have increased odds of experiencing client-perpetrated violence (27–29).

Secondly, FSWs who practiced mobile sex work were found to have increased odds of experiencing client-perpetrated GBV. The literature on the relationship between sex work-related mobility and exposure to client-perpetrated GBV are scarce. Moreover, to the best of our knowledge, this is the first study on the relationship between client-perpetrated GBV and sex work-related mobility in post-conflict settings. However, our finding is consistent with previous studies reporting that mobile FSWs were at increased odds of client-perpetrated GBV (9,10,30). This is because mobile FSWs tend to have less control over their work environment and many of the male clients of FSWs tend to refuse to use condoms with the new coming mobile FSWs (30). Therefore, to reduce client-perpetrated GBV, health programmers need to organize GBV interventions that reach all the regional hot-spots where the mobile FSWs operate.

Thirdly, our findings showed that FSWs living with HIV were at increased odds of reporting client-perpetrated GBV. This is in agreement with previous studies showing that living with HIV in the general population (6) as well as among the FSWs (2,31) increases a woman’s odds of experiencing GBV. The relationship between HIV and client-perpetrated GBV could have been mediated through accidental exposure of HIV status to clients, HIV stigma, and depression which are common in this population. However, it’s worth noting that the relationship between GBV and HIV is bidirectional because studies have indicated that people who experienced GBV were more likely to get HIV infection (7,32,33).

Lastly, client-perpetrated GBV did not show any significant relationship with income at the bivariate level. However, at the multivariate level, client-perpetrated GBV showed a strong positive association with low monthly income. Specifically, FSWs earning less than 50^th^ percentile of the average monthly income of FSWs were almost three times more likely to report client-perpetrated GBV compared to the relatively higher-income earners. Only a few studies examined the relationship between client-perpetrated GBV and FSWs’ monthly income. In Tanzania, the income level did not significantly affect the occurrence of client-perpetrated GBV (10). However, in Northern Ethiopia low earning FSWs were more likely to suffer from client-perpetrated GBV (34).

This high magnitude of client-perpetrated GBV among FSWs calls for more attention and resources to address this public health threat to the mental and sexual reproductive health of FSWs. The government should enact favorable laws that protect the FSWs against GBV. Secondly, because client-perpetrated GBV among FSWs is multi-dimensional, interventions to prevent and treat GBV in this population should target the low earning FSWs who are mobile, street-based, illicit drug users, and HIV-positive.

### Strengths and limitations of the study

Our study had several strengths: unlike most previous studies among the FSWs that used the non-probability methods of participants’ selection, we utilized a simple random sampling technique to select our sample. Thus, our study participants were representative of the FSWs. However, this study had some limitations; the information collected may have been influenced by recall bias since we asked FSWs about their personal experience of client-perpetrated GBV since joining sex work. In addition, we implored some sensitive information relating to sex work that might have been difficult to answer. However, we interviewed the FSWs in such a way that we reduce any chance of information bias.

## Conclusion

Our findings revealed a high magnitude of client-perpetrated GBV among FSWs in Gulu district, Northern Uganda. Further results indicated that FSWs who were street-based, mobile, HIV-positive, and low-income earners had increase odds of client-perpetrated GBV. Therefore, to maximize interventional benefits, future public health interventions to address client-perpetrated GBV in this population should be integrated within the existing HIV programs and should target the mobile, street-based, and low income earning FSWs.

## Data Availability

The datasets used and analyzed during the current study are available from the corresponding author on reasonable request to

## List of abbreviation

AIDS: Acquired Immunodeficiency Syndrome
ART: Antiretroviral Therapy
aOR: Adjusted Odd Ratio
CI: Confidence Interval
FSWs: Female Sex Workers
GBV: Gender-Based Violence
HIV: Human Immunodeficiency Virus
TASO: The AIDS Support Organization
UGX: Ugandan Shillings
uOR: Unadjusted Odd Ratio.

## Ethics approval and consent to participate

We obtained ethical clearance for this study from the Makerere University School of Public Health Higher Degrees, Research, and Ethics Committee. Each participant provided written informed consent. We maintained participants’ privacy and confidentiality throughout the different processes of participant’s enrollment into the study and data collection and analysis.

## Consent for publication

Not applicable

## Acknowledgments

We acknowledge Prof. Christopher Garimoi Orach, Prof. Fred Nuwaha, and Assoc. Prof. Noah Kiwanuka for their insightful contributions to our study designs. We acknowledge The AIDS Support Organization (TASO)-TASO Gulu for the access to their up-to-date database of FSWs in Gulu district. We acknowledge Ms. Ajwang Brenda for her role as a research assistant for this study. Last but not least, we acknowledge all the study participants and the peers of FSWs whose ideas and expertise informed the design and the conduct of our study and for enabling the smooth conduct of this study.

## Contributorship

SO, conceived and designed the study, collected and entered data, conducted data analysis, interpreted the findings, and wrote the first draft of the manuscript. RN, AC, and NMT conceived the study, supported data analysis, interpretation of results, and critically reviewed the draft manuscript. All authors approved the final manuscript for publication.

